# Domain-specific LLM Development and Evaluation – A Case-study for Prostate Cancer

**DOI:** 10.1101/2024.03.15.24304362

**Authors:** Amara Tariq, Man Luo, Aisha Urooj, Avisha Das, Jiwoong Jeong, Shubham Trivedi, Bhavik Patel, Imon Banerjee

## Abstract

In this work, we present our strategy for developing domain-specific large language models which cover the vocabulary of the target domain and train on reliable sources of clinical information. Prostate cancer was chosen as a use-case for this study. We collected more than 1.8 million clinical notes and radiology and pathology reports for 15341 patients treated for prostate cancer in Mayo Clinic across three sites and outpatient clinics. In addition to domain-specific training data, we built domain-specific tokenizers and devised knowledge-guided training strategies for LLM development. During the self-supervised training, LLM was forced to predict domain-specific information by marking clinical terms using UMLS parser. We evaluated the model for downstream tasks of clinical information prediction and question answering using quantitative and user evaluation study to measure the accuracy, reliability and information completeness. We compared the domain-specific model against similarly sized general purpose model GPT-2 and a three-times larger domain specialized model. i.e., BioGPT. Our model outperformed GPT-2 on both tasks by a wide margin. Our model was also able to outperform BioGPT on clinical information prediction tasks and showed some advantages over BioGPT in question-answering tasks.

## Introduction

Prostate cancer is the most commonly diagnosed non-skin cancer among men in the United States with a 5-year survival rate of about 100% for local or regional cancer^1,2^. However, the diagnosis of prostate cancer is often followed by immediate decline in mental and physical health^3,4^. Limited knowledge regarding the disease, embarrassment around physical examination such as rectal exam and discussion of sexual symptoms with healthcare professionals, potentially exacerbated depending on the provider gender^5,6^, are known causes of poor outcome among older patients and/or those belonging ethnic minorities ^7–9^. These uncertainties and anxiety related to sexual symptoms could lead patients to search for information through online search engines in which the patient may end-up with either incomplete, contradictory, misleading, and/or inaccurate information^10^. This, in turn, could cause further delay in treatment and poor outcomes.

During diagnosis, treatment planning, and post-treatment disease management, vast amounts of clinical information is generated and recorded by the care team about prostate cancer patients. This is usually in free-text form including clinical notes and radiology and pathology reports. Current developments in the field of large language modeling provide a unique opportunity to process this information for developing reliable agents for interactive knowledge sharing in a discreet way^11–13^. However, we hypothesize that general purpose LLMs, like GPT, are at a disadvantage when processing this information as they have not been trained for any specific domain. They may generate generic responses to domain-specific questions. They also suffer from hallucination, which can be especially dangerous in the sensitive domains like clinical medicine^13–16^. Research effort has been put into domain-specific LLM development such as MedPALM^17^ (540B), MedAlpaca^18^ (13B), and BioGPT^19^ (1.5B). However, these models are also broad and intended to cover complete domains of medicine and biology. To cover such vast fields, these models are growing in sizes reaching on the order of 100 billions of parameters. Medicine includes subspecialties, with different domain specific knowledge such as oncology, preventive medicine, and emergency medicine where the language and treatments differ.

Large text corpora are generally used to train large models. GPT^20^ and LLaMA^21^ have been trained on text crawled from the world wide web without verifying the source reliability. Training of specialized LLMs like BioGPT and MedPALM have been focused on domain-specific knowledge corpora like millions of scientific abstracts from PubMed. However, patient records are arguably the most critical source of information to capture the domain of any disease. The records include several distinct documents including oncology notes, nursing notes, ED reports, discharge summaries, and radiology and pathology reports. While GPT2 and LLaMA, and even medicine-focused BioGPT, have been trained on extremely large amounts of information, their training data cannot include such patient-specific documents because of patient privacy requirements set by Health Insurance Portability and Accountability Act (HIPAA). We argue that this puts them at an inherent disadvantage in terms of understanding the clinical decision making process and information dissemination to the patients themselves. While research papers and expert treatment guidelines can impart knowledge about latest treatment options and their general pros and cons, clinical notes actually describe how physicians and patients may decide on a treatment option and their following outcomes. We argue that inclusion of clinical notes and reports is critical when building a language understanding and generation tool for any particular disease.

We present our work on development of a domain-specific LLM to overcome the shortcomings of general purpose LLMs when applied to sensitive domains like medical decision making and information dissemination. We chose prostate cancer as our specific use case. We chose a relatively smaller size for our LLM (i..e, 124M parameters) and collected and parsed prostate cancer related patients’ data including clinical notes and radiology and pathology reports for its training. We also devised domain-specific self-supervised training techniques to ensure that model learned the nuances of the chosen domain. We evaluated the proposed LLM against similarly sized general purpose LLM (GPT2-small) as well as larger domain-specific LLM (BioGPT) on two tasks; i) clinical information mask prediction, and ii) question-answering. We also measured the performance in a user study with 3 independent raters who evaluated the generated responses in terms of completeness, correctness and relevance.

## Methodology

In this section, we describe several steps involved in the development and training of a domain-specific LLM. We argue that these steps form a framework that could be replicated to other domains like breast cancer. Figure 1 shows the overall framework for domain-specific LLM development and application.

**Figure 1:**
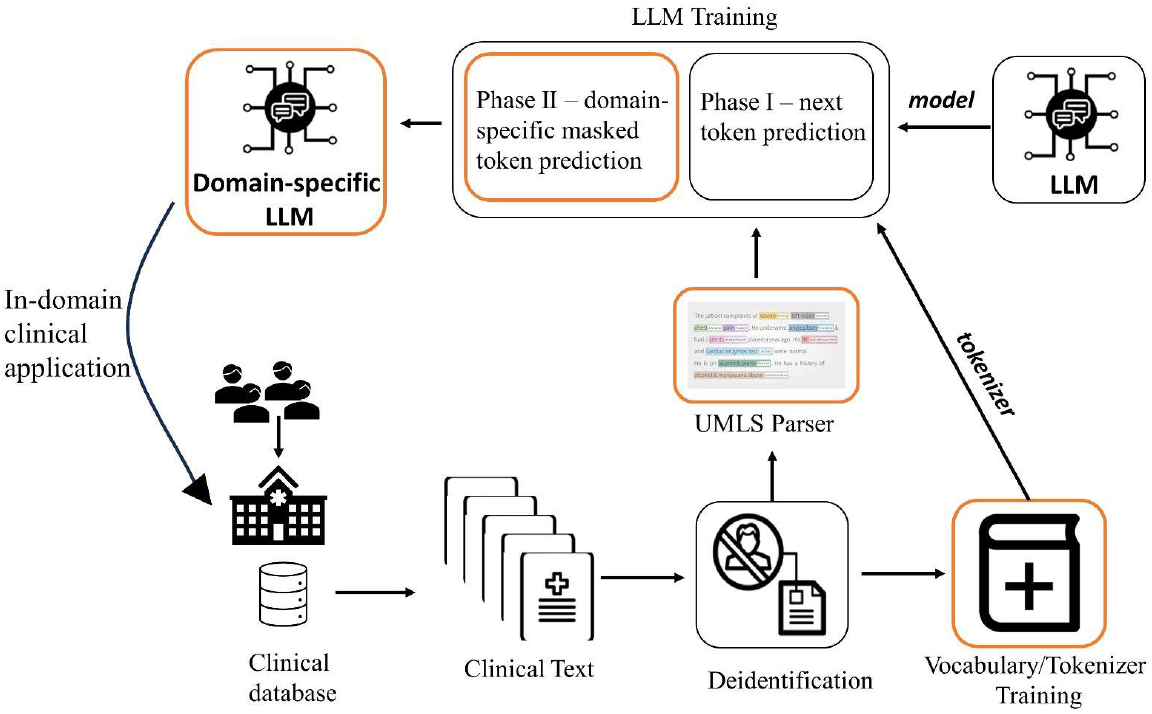
Domain-specific LLM training and application framework; Highlighted (orange-colored) modules indicate domain-specific modifications to LLM design framework.

### Data curation

With the approval of the internal review board (IRB) at Mayo Clinic, we collected clinical notes concerning 23,665 patients treated for prostate cancer at all three major sites of Mayo Clinic (Rochetser, Arizona, Florida). Statistics of these patients are provided in Table 1. This data included not only all clinical notes and radiology and pathology reports. We developed rule-based NLP techniques to gather information about cancer characteristics of these patients such as lesion size and Gleason score. These patients were also part of an enterprise-wide cancer registry which recorded their clinical outcome information.

**Table 1:** Cohort characteristics for the prostate cancer patients. Earliest inclusion date is 2017, thus the followup is limited.

| Characteristics | Value |
| --- | --- |
| Patients | 15,341 |
| Total number of clinic notes | 1,852,213 |
| Radiology reports | 23,665 |
| Followup | mean: 2.8+/-1.8 years<br>max: 6.6 years |
| Age | 66.7+/-8.6 years |
| Race | Caucasian: 14,213 (93%)<br>African American: 548 (4%)<br>Asian: 171 (1%) |
| Gleason Score | 6 or lower: 17%; 7 or higher: 65% |
| PSA | PSA<=10: 57%; 10<PSA<20: 13%; PSA>=20:10% |
| Tumor size | mean: 366.3 mm, min: 1.0mm, max: 990 mm |

### Text Parsing

Clinical data collected for the patients cohort included ∼ 1.8 million clinical notes (including oncology and all other notes) and 23665 radiology and pathology notes. This data was used for training of our domain-specific LLM after several steps of parsing and cleaning.

#### Anonymization

Clinical notes contain not only important clinical information about patient-provider interaction, they also contain identifiable information about both patients and providers. While this data was not intended for public release, we wanted to ensure patient and provider privacy was not violated even by sharing the weights of our domain specific language model. Large language models display behaviors like memorization which leaves open the possibility of unmasking identifiable patient information used during the training phase. Therefore, we anonymized all clinical notes prior to their use in training of the proposed LLM using a pretrained BiLSM model with CRF output layer ^22^. Information like patient and provider names, dates, location were replaced with special tokens like “[NAME]”, “[DATE]”, and “[LOC]”. Only imaging finding sections from radiology and pathology information were used for LLM training. These sections are devoid of any patient information. Electronic signatures of providers were dropped using regular expression to ensure provider privacy.

#### Filtering

Textual dataset cleaning steps included filtering of very short (< 3 words) and very long sentences (>100 words). Given the repetition practice, we also used fuzzy matching techniques to filter out sentences which are too similar to other sentences within the clinical notes for one patient. This is done to improve the quality of training data. Repetition of standard/templated sentences may skew the behavior of the trained LLM towards generating such templated sentences at inference time.

#### Clinical Information Marking

We wanted to ensure that domain-specific LLM was focused on cancer treatment, side-effects, outcomes and symptoms related information during the training phase. To achieve this, we first needed to identify domain-specific information from free-text clinical notes. We used the Unified Medical Language System (UMLS) metathesaurus made available by the National Institute of Health^23^. UMLS framework is designed to standardize medical vocabularies and ontologies. Text parsing tools designed on top of UMLS identify medical concepts from free-text and map them to unique identifiers (CUI). Concepts are categorized into 127 “semtypes”. We selected 36 semtypes relevant to treatment, symptoms, side effects, clinical status and anatomical details. Filtered text from clinical notes was divided into sentences and passed through UMLS parser one by one and relevant medical entities and their location in the sentences were stored. We used a python parser for UMLS designed on a type of medical named entity extractor for parsing^24^. Our processing indicated that 84% of the sentences from clinical notes included important medical concepts. Medical concepts have varying distribution in the data with 33% concepts with one occurrence and 0.3% with greater than 10000 occurrences. A total of 193,936 concepts were identified. We argue that extremely common terms (>10000) may act as stop words such that they have no distinctive capabilities to establish differences between different text samples. For example, the concept “prostate cancer”,, “lesion”, “prostate specific antigen”, “elevated PSA” would have little distinctive capabilities within a corpus of all prostate cancer patients.

### Domain-specific Tokenizer

Modern LLMs rely on word-piece tokenizers that meet the challenge of out-of-vocabulary token by chopping up the word into smaller foundational pieces such as “carcinoma” may be chopped into “car_” and “_cinoma”. However, we argue that chopping up important domain specific terms into multiple tokens may hinder model’s focus on learning about the whole clinical terms and their context. A passage full of relevant medical information containing N words may be divided in M tokens when M>>N. In extreme cases, M may exceed the length of the context window causing the model to ignore some information. On the other hand, a domain-specific tokenizer may be able to preserve boundaries of medical concepts and entities, thus shortening the length of the relevant text and allowing for better attention within the textual context. Therefore, we decided to train a domain-specific wordpiece tokenizer with vocab size of 50 thousand which was similar in size to popular tokenizers such GPT-2 tokenizer (vocab size of 50257). The tokenizer was trained on filtered clinical text as described in the previous section.

### Domain-specific LLM

We based the architecture of our LLM on popular 12-layer masked self-attention decoder based architecture similar to the one used by GPT-2^20^. The model was composed of about 124M trainable parameters. LLM training on free-text in self-supervised fashion is a critical component of LLM development. Simple language based self-supervision pre-training tasks like the prediction of the next token seem enough for general purpose models like GPT to gain fundamental understanding of natural language. We built upon this line of research by introducing domain-specific self-supervision in addition to generic self-supervision. Our LLM was trained in two phases.

#### Phase I - General purpose language understanding

Under this phase, our LLM was trained on free text data of prostate cancer patients including clinical notes and radiology and pathology reports for the task of next token prediction. Token sequences for free-text were generated using our domain-specific tokenizer. We named the model trained under this task only as PCa-LLM.

#### Phase II - Domain specific language understanding

In phase II of training, we aimed for the model to learn clinical language specific to prostate cancer. As described earlier, we marked clinical terms in free-text data of prostate cancer patients using UMLS parser. In this phase of training, we used masked clinical token prediction as a self-supervised training task.

Each text snippet may contain more than one clinical term. Terms were chosen with 50% probability for masking. Note that one term may be split into multiple tokens. In our training scheme, all tokens belonging to that term were masked and the model was trained to predict masked tokens.

For example, the sentence “*common side effects of hormone therapy may include shrinkage of testicles*” included clinical terms of “hormone therapy” and “shrinkage of testicles”. If “hormone therapy” was selected randomly for marking, all tokens belonging to it ({“hormone”, “therapy”}) would be masked. In this case, the model received “*common side effects of <mask> <mask> may include shrinkage of testicles*” and was tasked with predicting “hormone” and “therapy”.

We argue that this training scheme allowed the domain-specific LLM to focus on domain-specific language in addition to learning correlation between treatments, side effects, symptoms, and cancer characteristics. Such correlations can be essential for domain specific downstream tasks like patient education or treatment recommendation. For such downstream tasks, the model must be able to predict clinical terms using the context provided in the input text. This phase of training essentially taught the model to do the same. We call the model that underwent both phases of training PCa-MLM.

### Evaluation Tasks

After training, we evaluated the model on two downstream tasks; i) masked clinical information prediction, and ii) question-answering.

#### Masked clinical information prediction

This task was similar to phase II of training. We randomly selected clinical terms and masked them in a set of held-out sentences (text not used during the training phase). The model was tasked with retrieving masked terms from the vocabulary of its tokenizer. We evaluated the model through an evaluation metric for information retrieval systems, i.e., recall@K where K was varied between 1 and 10, based on ranked retrieval results generated by the model probability.

#### Clinical question-answering

One of the most valuable downstream applications for any LLM can be a question-answering framework that can allow the users to interact with the LLM in an interactive and flexible manner. With this in mind, we chose question-answering as the second evaluation task.

We manually curated questions from treatment guidelines set by the American Cancer Society for prostate cancer. We browsed through each section of the guidelines and defined question-answer pairs of the following broad categories.

- Treatment path recommended for cancer at different stages/risk category
- Treatment path recommended for cancer at different stages/risk category under certain conditions (older patient, patients in poor health, patients with certain comorbidities)
- Side effects of treatment
- Long term risks of treatment
- Medication used in treatment
- Comparative pros and cons of different treatment

About 300 question answer pairs were manually curated. We then employed open-source general purpose LLM, i.e., LLaMA (13B)^21^ within a generative framework to paraphrase each question 4 different ways. After filtering for nonsensical paraphrasing such as empty strings, we were left with 1,222 QA pairs. Randomly selected unique 35 QA pairs were kept in a holdout set for evaluation purposes after dropping paraphrases of the same questions.

## Results

In this section, we describe the results of evaluative experiments. We selected two models as comparative baselines for our model, i.e., GPT2-small and BioGPT. Selected version of GPT2 was similarly sized as our model but lacked the advantages of domain-specific vocabulary, training data, and training tasks. BioGPT was about three times bigger and was designed to cover a vast domain of medicine as a whole, and thus, had domain-specific vocabulary. Comparison with this model was meant to establish benefits of fine-grained domain definition (cancer vs. generic medicine).

### Domain vocabulary Coverage

We applied general purpose GPT-2 tokenizer, medicine focused BioGPT tokenizer, and our tokenizer on clinical terms extracted by UMLS parser. GPT2 tokenizer tended to chop up medical concepts like “prostatectomy” (GPT2 tokenizer: 2 tokens, Our tokenizer: 1 token), “diabetes mellitus” (GPT2 tokenizer: 4 tokens, Our tokenizer: 2 token), etc. BioGPT tokenizer showed better clinical vocabulary coverage for up to 3 tokens than our tokenizer. However, it still chopped prostate cancer related words like “genitourinary” (GPT2: 5 tokens, BioGPT: 5 tokens, Ours: 1 token) and “erectile dysfunction” (GPT2: 5 tokens, BioGPT: 3 tokens, Ours: 2 token). Figure 2 shows distribution of medical concepts chopped into token sets of varying length by the three different tokenizers. Even though GPT2 coverage is minimal for the clinical terms with lowest number of tokens <=1, BioGPT and our model have similar coverage of the medical terms.

**Figure 2:**
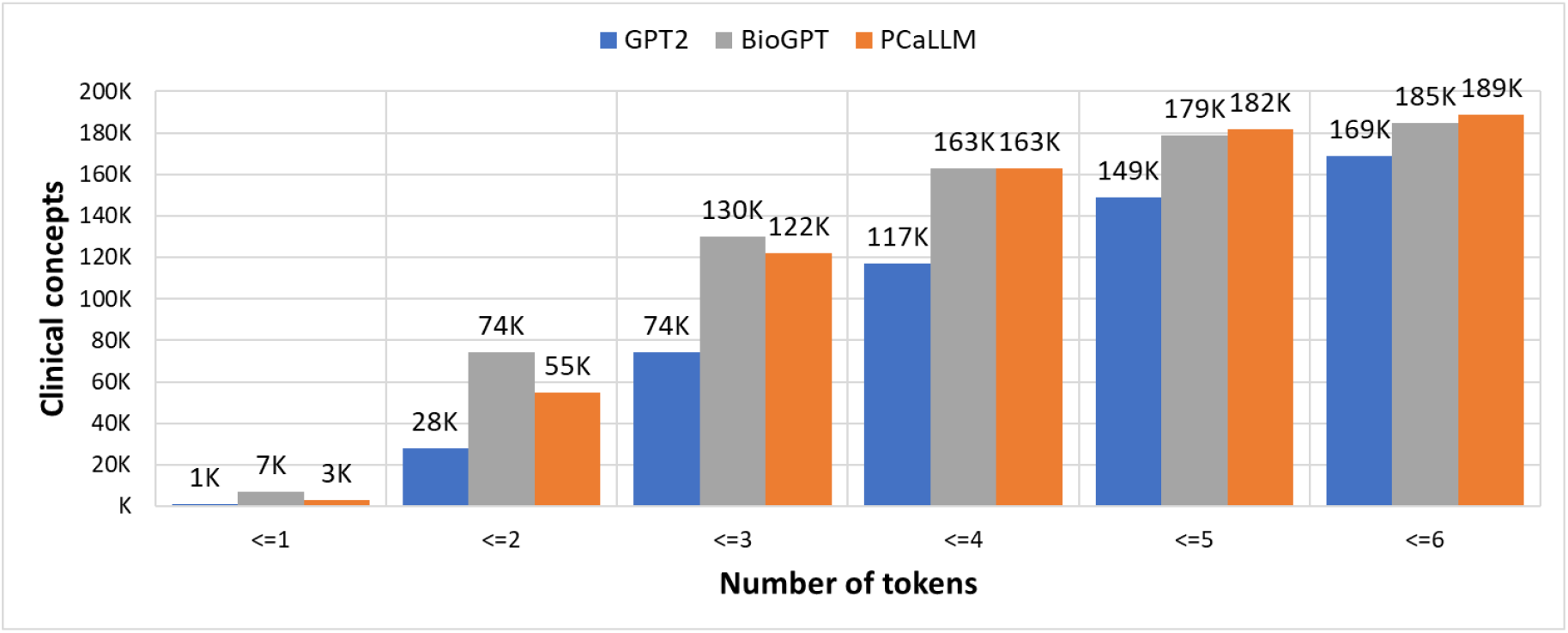
Distribution of clinical concepts across number of tokens formed by three different tokenizers

### Masked clinical information prediction

For masked clinical information retrieval, each model produced a probability estimate for every tokens in its vocabulary for prediction as replacement for the masked token. Hence, a ranked list of all tokens in vocabulary (sorted in descending order of probability estimate) was available for each masked token. Each model was evaluated to see if the correct token was found in top K tokens for each masked token (recall@K). The performance was aggregated for all masked tokens (Table 2). We evaluated two phases of our model, i.e., *Phase I:* PCa-LLM and *Phase II:* PCa-MLM. Both versions significantly outperformed both comparative models with significant margin at each K, even when the vocabulary coverage for BioGPT is similar to the domain specific model.

**Table 2:** Clinical information retrieval performance for different LLMs and proposed domain specific model. Phase I training -PCa-LLM and phase II training PCa-MLM.

| Model | Recall@10 | Recall@5 | Recall@1 |
| --- | --- | --- | --- |
| GPT-2 | 0.017 | 0.010 | ~0.000 |
| BioGPT | 0.017 | 0.010 | 0.003 |
| PCa-LLM | 0.228 | 0.173 | 0.076 |
| PCa-MLM | **0.305** | **0.239** | **0.111** |

### Question-answering

For evaluation of the question-answering task, we relied on a user study where 3 independent users were asked to evaluate the responses of each model on the basis of three criteria.

#### Correctness

if the generated response was correct according to the clinical guidelines; the response did not have to be complete or cover all aspect of ground truth answer;

#### Completeness

if the generated response was complete, covered all aspects of the groundtruth answer;

#### Relevance

if the generated response was relevant to the question; response may be incorrect or incomplete based on the clinical guideline;

While correctness and completeness evaluate the accuracy of the response, the third criteria was introduced to assess the model’s comprehension of the question. Models, especially general purpose models - GPT2, may generate generic responses if they are unable to understand the granularity of the question.

After de-identification of the model names, all users were asked to provide 1/0 response for each question for all criteria; 1 indicated that the criteria was met (complete, correct or relevant) while 0 indicated that the generated response did not meet the criteria (incomplete, incorrect, or irrelevant). Responses in each category by every user were summed over 35 questions. By aggregate opinion of users (median scores), our model outperformed both comparative models. Table 3 shows the results from our user study where the median value for all the criteria are rated higher for our model. User 3 considers BioGPT responses rated higher for correctness and completeness while still rating our model higher for relevance based on the generated responses being more relevant for the prostate cancer domain. Higher numbers indicate better performance. Table 4 shows sample answers generated by all three models.

**Table 3:** User evaluation study for question-answering performance of different LLMs. Represents the count and (%).

| Model | Criterion | user 1 | user 2 | user 3 | Median |
| --- | --- | --- | --- | --- | --- |
| GPT2 | **Correctness** | 16 (46%) | 7 (20%) | 4 (11%) | 7 (20%) |
|  | **Completeness** | 12 (34%) | 6 (17%) | 3 (9%) | 6 (17%) |
|  | **Relevance** | 31 (89%) | 17 (49%) | 9 (26%) | 17 (49%) |
| BioGPT | **Correctness** | 23 (66%) | 21 (60%) | **23 (66%)** | 23 (66%) |
|  | **Completeness** | 14 (40%) | 20 (57%) | **14 (40%)** | 14 (40%) |
|  | **Relevance** | 35 (100%) | 26 (74%) | 27 (77%) | 27 (77%) |
| PCa-MLM | **Correctness** | **27 (77%)** | **27 (77%)** | 18 (51%) | **27 (77%)** |
|  | **Completeness** | **14 (40%)** | **23 (66%)** | 11 (31%) | **14 (40%)** |
|  | **Relevance** | **35 (100%)** | **32 (91%)** | **29 (83%)** | **32 (91%)** |

**Table 4:** Sample answers generated by different LLMs for the same questions related to prostate cancer.

|  |  |
| --- | --- |
| **Question 1** | **What treatment is recommended if the PSA level is greater than 20 and prostate cancer with a Gleason score of 8 to 10 is found in lymph nodes?** |
| *Groundtruth Answer* | hormone therapy, possibly along with external radiation |
| *GPT-2 response* | it means that when people are given drugs that are similar to what they are given in clinical trials, |
| *BioGPT response* | hormone therapy, possibly along with external radiation therapy, may be recommended. This is a type of focal therapy |
| *PCaMLM response* | hormone therapy, possibly along with external radiation therapy |
| **Question 2** | **What is ablative treatment?** |
| *Groundtruth Answer* | heat or cold or other methods to destroy (ablate) prostate tissue, rather than removing it with surgery or treating it with radiation. Methods include high intensity focused ultrasound (HIFU) and cryotherapy. |
| *GPT-2 response* | surgery (TURP) may be recommended to relieve symptoms like bleeding and urinary obstruction |
| *BioGPT response* | Cryotherapy (also called cryosurgery or cryoablation) is the use of very cold temperatures to freeze |
| *PCaMLM response* | HIFU uses highly focused ultrasound beams to heat and destroy prostate tissue and shrink prostate over time |
| **Question 3** | **What are the side effects of bisphosphonates** |
| *Groundtruth Answer* | flu-like symptoms and bone or joint pain, kidney problems, so patients with poor kidney function might not be able to be treated with these medicines |
| *GPT-2 response* | Corticosteroids are drugs that target PD-1, a checkpoint protein on immune system cells |
| *BioGPT response* | denosumab (Xgeva), a bisphosphonate-like zoled |
| *PCaMLM response* | pain, swelling in the legs or feet, nausea, and diarrhea. These drugs can also cause kidney damage. |
| **Question 4** | **What are common side effects of chemotherapy drugs?** |
| *Groundtruth Answer* | Hair loss, Mouth sores, Loss of appetite, Nausea and vomiting, Diarrhea, Increased chance of infections (from having too few white blood cells), Easy bruising or bleeding (from having too few blood platelets), Fatigue (from having too few red blood cells) |
| *GPT-2 response* | doctor might recommend external radiation therapy to the prostate bed (the area where the prostate was before it was removed), |
| *BioGPT response* | allergic reaction, nerve damage, leukemia, and increased risk of blood clots in the legs or lungs. |
| *PCaMLM response* | fatigue, nausea, vomiting, and diarrhea. |
| **Question 5** | **What are common side effects of brachytherapy?** |
| *Groundtruth Answer* | bowel problems, urinary problems, erection problems , |
| *GPT-2 response* | nausea, diarrhea, and feeling weak or tired. |
| *BioGPT response* | (1) bowel problems, (2) urinary problems, (3) erection problems |
| *PCaMLM response* | Pain in the treatment area, Blood in the urine, An increased risk of a urinary tract infection (UTI) |

## Discussion

Even though general purpose huge LLMs have demonstrated amazing capabilities to generalize to a wide variety of domains as zero-shot frameworks, their performance may still suffer in terms of relevance or specificity to a sensitive domain and may “*hallucinate*” while being forced to generate critical information^14,16,25^. Training of these models require humongous text corpora usually curated from publicly available text context available on the world wide web. Privacy concerns make it impossible to train such models in private data which may be critical for medicine related fields. We present a framework for *design and development of domain-specific LLM with prostate cancer chosen as a use-case*. Our framework includes domain-specific customization steps for tokenizer design, data curation, and self-supervised two-phases training. We argue that development of domain-specific LLMs provides a systematic solution to the challenges faced in terms of deployment of huge general purpose LLMs in sensitive domains. While some research has been done to specialize LLMs for sensitive domains like medicine and biology, this line of research usually focused on curation of large domain-specific datasets^17,19^. BioGPT and MedPALM are two popular biomedical LLMs developed to support this line of research. These LLMs still cover vast fields of medicine and biology and their training has been focused on public datasets. These LLMs have absorbed the knowledge of vast sources of medicine related information but have had little exposure to the real clinical data and thus patient-specific treatment decisions. In comparison, our framework focuses on training the model on patient EHR data collected by healthcare institutions including clinical notes and radiology and pathology reports. Thus, the models trained under our framework will be exposed to unique characteristics of patient-physician exchange for the particular targeted domain and able to learn the nuance of the treatment management.

We trained a small sized LLM (124M parameters) for the particular use case of prostate cancer which not only outperformed similarly sized general purpose LLM (GPT-2 small) but also outperformed the three times larger specialized LLM (BioGPT) for two downstream tasks. Specialized tokenizer allows the LLM to understand and focus on domain-specific terms instead of chopping them up into small generic tokens (see Figure 2). GPT2 tokenizer chops up clinical terms in a larger number of tokens while our specialized tokenizer and BioGPT tokenizer can comprehend most of the clinical terms as a whole. As these tokenizers were designed for the fields of medicine and biology, it tends to describe medicine related terms in fewer tokens. It covers more clinical terms within 3 tokens splitting than or tokenizer. However, clinical terms which are more specific to prostate cancer(i.e., found more commonly in prostate cancer related clinical text) are better handled by our tokenizer than BioGPT tokenizer. These terms include “erectile dysfunction”, “primary malignant neoplasm”, “prostatic hypertrophy”, “cystolitholapaxy”, “cystourethroscopy”, and others. In addition, several prostate cancer related drug names are part of our specialized vocabulary including “abiraterone” and “docetaxel”.

For the task of masked clinical term prediction, it is intuitive that PCa-MLM (phase I and II of training) outperformed PCa-LLM (phase I training only) as phase II training was performed using a very similar self-supervision task. However, even PCa-LLM outperformed GPT-2 and BioGPT by wide margins which has been trained using generic self-supervision tasks of next token prediction. This speaks to the benefits of specialized domain-specific tokenizers. BioGPT tokenizer displayed somewhat better coverage of generic clinical vocabulary than our tokenizer (as shown in Figure 2) but the model still performed extremely poorly for prediction of domain-specific (prostate cancer related) clinical information retrieval.

One of the main applications of our specialized LLM can be patient education as an interactive question answering framework. Therefore, we tested the performance of the model on a question-answering task. Note that these questions were curated from the website of the *American Cancer Society*. Since this data is publicly available, it is likely to be included in the training data of GPT2 and BioGPT’s training corpus. On the other hand, our model was only trained on clinical data generated within the hospital. It was never trained explicitly on the textual guidelines. Despite this, our model outperformed GPT2 by a wide margin on all three evaluation criteria. Table 4 shows some interesting examples of the response. It is clear that GPT2 often generated generic or irrelevant responses. Even when answering questions about side effects (Question 5), it described generic side effects like “feeling weak and tired” which was not very specific to the treatment in question. BioGPT does not suffer from this problem. Its answers were mostly related to the question even when it failed to produce the correct answer. For example, it generated names of bisphosphonate drugs when the question was about side effects of bisphosphonates (question 3). In answer to question 1, it generated a treatment option which is available for cancer (focal therapy) but was not part of the ground truth answer. PCaMLM and BioGPT responses differed from each other for question 2 and 4, but were still relevant and somewhat appropriate for the question.

Our experiments clearly establish the benefits of developing and employing domain-specific LLMs for sensitive domains like medicine and healthcare, especially for building interactive knowledge dissemination tools. We plan to expand upon this line of research by developing patient and physician facing chatbots on top of our domain specific LLM for the use case of prostate cancer. While it will be challenging to curate training datasets, i.e., question-answer pairs, manually, we plan to build upon automatic question-answer generation frameworks^26^. This line of research will provide a solution to reliability and specificity issues faced by general purpose chatbot like ChatGPT in the field of medicine.

## Data Availability

All data produced in the present study are available upon reasonable request to the authors

## References

1. Brawley OW. Trends in prostate cancer in the United States. Journal of the National Cancer Institute Monographs. 2012;2012(45):152–156.

2. Henley SJ, Ward EM, Scott S, et al. Annual report to the nation on the status of cancer, part I: National cancer statistics. Cancer. 2020;126(10):2225–2249.

3. Petros NG, Alvarsson-Hjort J, Hadlaczky G, et al. Predictors of the Use of a Mental Health–Focused eHealth System in Patients With Breast and Prostate Cancer: Bayesian Structural Equation Modeling Analysis of a Prospective Study. JMIR cancer. 2023;9:e49775.

4. Crump C, Stattin P, Brooks JD, et al. Risks of alcohol and drug use disorders in prostate cancer survivors: a national cohort study. JNCI Cancer Spectrum. 2023;7(4):pkad046.

5. Fink M, Klein K, Sayers K, et al. Objective data reveals gender preferences for patients’ primary care physician. Journal of Primary Care & Community Health. 2020;11:2150132720967221.

6. Steinkohl F, Luger AK, Gruber L, et al. Acceptance of female urologists among patients with suspected prostate disease. Translational Andrology and Urology. 2021;10(7):2938.

7. Ravi P, Karakiewicz PI, Roghmann F, et al. Mental health outcomes in elderly men with prostate cancer. In: Vol 32. Elsevier; 2014:1333–1340.

8. Shaw B, Walter FM, Hamilton W, Martins T. Symptom appraisal and help seeking in males with symptoms of possible prostate cancer: a qualitative study with an ethnically diverse sample in London. British Journal of General Practice. 2023;73(732):e502–e510.

9. Wray RJ, McClure S, Vijaykumar S, et al. Changing the conversation about prostate cancer among African Americans: results of formative research. Ethnicity & health. 2009;14(1):27–43.

10. Muzii B, Di Bello F, Carraturo F, et al. Mental Health of Prostate Cancer Patients: Content Review on YouTubeTM. International Journal of Environmental Research and Public Health. 2023;20(6):4721.

11. Harris E. Large language models answer medical questions accurately, but can’t match clinicians’ knowledge. JAMA. Published online 2023.

12. Omiye JA, Gui H, Rezaei SJ, Zou J, Daneshjou R. Large Language Models in Medicine: The Potentials and Pitfalls: A Narrative Review. Annals of Internal Medicine. 2024;177(2):210–220.

13. Thirunavukarasu AJ, Ting DSJ, Elangovan K, Gutierrez L, Tan TF, Ting DSW. Large language models in medicine. Nature medicine. 2023;29(8):1930–1940.

14. Alkaissi H, McFarlane SI. Artificial hallucinations in ChatGPT: implications in scientific writing. Cureus. 2023;15(2).

15. Chen S, Kann BH, Foote MB, et al. Use of artificial intelligence chatbots for cancer treatment information. JAMA oncology. 2023;9(10):1459–1462.

16. Sallam M. ChatGPT utility in healthcare education, research, and practice: systematic review on the promising perspectives and valid concerns. In: Vol 11. MDPI; 2023:887.

17. Singhal K, Azizi S, Tu T, et al. Large language models encode clinical knowledge. Nature. 2023;620(7972):172–180.

18. Han T, Adams LC, Papaioannou JM, et al. MedAlpaca--an open-source collection of medical conversational AI models and training data. arXiv preprint arXiv:230408247. Published online 2023.

19. Luo R, Sun L, Xia Y, et al. BioGPT: generative pre-trained transformer for biomedical text generation and mining. Briefings in bioinformatics. 2022;23(6):bbac409.

20. Radford A, Wu J, Child R, Luan D, Amodei D, Sutskever I. Language models are unsupervised multitask learners. OpenAI blog. 2019;1(8):9.

21. Touvron H, Lavril T, Izacard G, et al. Llama: Open and efficient foundation language models. arXiv preprint arXiv:230213971. Published online 2023.

22. Trienes J, Trieschnigg D, Seifert C, Hiemstra D. Comparing rule-based, feature-based and deep neural methods for de-identification of dutch medical records. arXiv preprint arXiv:200105714. Published online 2020.

23. Bodenreider O. The unified medical language system (UMLS): integrating biomedical terminology. Nucleic acids research. 200432 (suppl_1):D267–D270.

24. Soldaini L, Goharian N. Quickumls: a fast, unsupervised approach for medical concept extraction. In: ; 2016:1–4.

25. Kojima T, Gu SS, Reid M, Matsuo Y, Iwasawa Y. Large language models are zero-shot reasoners. Advances in neural information processing systems. 2022;35:22199–22213.

26. Luo M, Mitra A, Gokhale T, Baral C. Improving biomedical information retrieval with neural retrievers. In: Vol 36. ; 2022:11038–11046.

